# Higher Temperature, Pressure, and Ultraviolet Are Associated with Less COVID-19 Prevalence – Meta-Regression of Japanese Prefectural Data

**DOI:** 10.1101/2020.05.09.20096321

**Authors:** Hisato Takagi, Toshiki Kuno, Yujiro Yokoyama, Hiroki Ueyama, Takuya Matsushiro, Yosuke Hari, Tomo Ando

## Abstract

A recent study from China suggests that high temperature and ultraviolet (UV) radiation cannot decrease the epidemics of Coronavirus disease 2019 (COVID-19). To determine whether COVID-19 prevalence is modulated by meteorological conditions, meta-regression of Japanese prefectural data was herein conducted. We extracted integrated number of patients testing positive for COVID-19 in each Japanese prefecture on 18 May 2020, population per 1-km^2^ inhabitable area in the prefecture in 2020, and monthly meteorological conditions at each prefectural capital city for 4 months (from January to April 2020). We averaged or cumulated the monthly data for the 4 months. To adjust for prefectural population density, we defined the COVID-19 prevalence as the integrated number of patients testing positive divided by the population per 100-km^2^ inhabitable area. Random-effects meta-regression was performed. A slope of the meta-regression line was significantly negative for mean air temperature (coefficient, −0.134; *P* = 0.019), mean sea level air pressure (−0.351; *P* = 0.001), and mean daily maximum UV index (−0.908; *P* = 0.012), which indicated that COVID-19 prevalence decreased significantly as air temperature, air pressure, and UV index increased. In conclusion, higher temperature, pressure, and UV may be associated with less COVID-19 prevalence, which should be confirmed by further epidemiological investigations taking other risk and protective factors of COVID-19 into account.

## Introduction

In Northern Europe, low air temperature and ultraviolet (UV) index have been reported as the most important meteorological predictors for the transmission of influenza virus.^1^ Whereas, a recent study from China suggests that high temperature and UV radiation cannot decrease the epidemics of Coronavirus disease 2019 (COVID-19).^2^ To determine whether COVID-19 prevalence is modulated by meteorological conditions, meta-regression of Japanese prefectural data was herein conducted.

## Methods

We extracted 1) integrated number of patients testing positive for COVID-19 in each Japanese prefecture on 18 May 2020 from the Ministry of Health, Labour and Welfare (https://www.mhlw.go.jp/content/10906000/000631885.pdf), 2) population per 1-km^2^ inhabitable area in the prefecture in 2020 from the Statistics of Japan (https://www.e-stat.go.jp/stat-search?page=1&toukei=00200502), and 3) monthly meteorological conditions at each prefectural capital city for 4 months (from January to April 2020) from the Japan Meteorological Agency (https://www.data.jma.go.jp/obd/stats/data/en/smp/index.html, https://www.data.jma.go.jp/gmd/env/uvhp/link_uvindex_month54.html) (Table 1). The meteorological conditions included 1) mean of air temperature (C), wind speed (m/s), sea level air pressure (hPa), relative humidity (%), possible sunshine (%), and daily maximum UV index; and 2) total of sunshine duration (h) and precipitation (mm). We averaged or cumulated the monthly data for the 4 months (UV index was available for 3 months, from January to March 2020). To adjust for prefectural population density, we defined the COVID-19 prevalence as the integrated number of patients testing positive divided by the population per 100-km^2^ inhabitable area. Random-effects meta-regression was performed using OpenMetaAnalyst (http://www.cebm.brown.edu/openmeta/index.html). A meta-regression graph depicted the COVID-19 prevalence (plotted as the logarithm transformed prevalence on the y-axis) as a function of a given factor (plotted as a meteorological condition on the x-axis).

**Table 1.** Extracted data in each Japanese prefecture

| Prefecture | Prefectural capital city | Patients testing positive for COVID-19 (*N*) | Population/100-km<sup>2</sup> inhabitable area (*N*) | COVID-19 prevalence (/population/100-km<sup>2</sup> inhabitable area) | Mean air temperature (C) | Mean wind speed (m/s) | Mean sea level air pressure (hPa) | Mean relative humidity (%) | Mean possible sunshine (%) | Total sunshine duration (h) | Total precipitation (mm) | Mean daily maximum UV index |
| --- | --- | --- | --- | --- | --- | --- | --- | --- | --- | --- | --- | --- |
| Tokyo | Tokyo | 5065 | 972400 | 0.005209 | 9.7 | 2.8 | 1015.4 | 63 | 53 | 731.2 | 577.5 | 2.4 |
| Osaka | Osaka | 1771 | 662270 | 0.002674 | 10.4 | 2.5 | 1018.1 | 60 | 48 | 684.2 | 342.5 | 2.5 |
| Kanagawa | Yokohama | 1276 | 623900 | 0.002045 | 10.3 | 3.8 | 1015.3 | 61 | 55 | 751.3 | 568.0 | 2.4 |
| Hokkaido | Sapporo | 1014 | 23630 | 0.042912 | 1.4 | 3.3 | 1014.6 | 69 | 40 | 554.9 | 368.5 | 1.6 |
| Saitama | Saitama | 991 | 283600 | 0.003494 | 9.0 | 2.6 | — | — | — | 785.0 | 373.5 | 2.4 |
| Chiba | Chiba | 900 | 175980 | 0.005114 | 10.3 | 3.6 | 1015.5 | 58 | 51 | 711.1 | 549.0 | 2.4 |
| Hyogo | Kobe | 700 | 197060 | 0.003552 | 10.5 | 3.8 | 1018.1 | 62 | 51 | 709.2 | 317.0 | 2.6 |
| Fukuoka | Fukuoka | 658 | 184930 | 0.003558 | 11.4 | 2.9 | 1019.4 | 65 | 45 | 638.9 | 459.0 | 2.5 |
| Aichi | Nagoya | 506 | 252220 | 0.002006 | 9.7 | 3.3 | 1017.1 | 63 | 56 | 786.8 | 369.5 | 2.5 |
| Kyoto | Kyoto | 358 | 220730 | 0.001622 | 9.5 | 2.1 | 1018.0 | 64 | 42 | 595.6 | 333.0 | 2.4 |
| Ishikawa | Kanazawa | 287 | 82120 | 0.003495 | 8.3 | 4.4 | 1017.8 | 66 | 35 | 507.6 | 697.5 | 2.1 |
| Toyama | Toyama | 227 | 56980 | 0.003984 | 7.7 | 3.1 | 1017.9 | 76 | 35 | 495.5 | 694.0 | 2.0 |
| Ibaragi | Mito | 168 | 72370 | 0.002321 | 8.1 | 2.5 | 1015.6 | 66 | 56 | 771.2 | 453.0 | 2.3 |
| Hiroshima | Hiroshima | 166 | 121890 | 0.001362 | 10.0 | 3.4 | 1018.9 | 57 | 51 | 716.0 | 471.0 | 2.5 |
| Gifu | Gifu | 150 | 90320 | 0.001661 | 9.6 | 2.8 | 1017.3 | 58 | 53 | 746.0 | 439.5 | 2.4 |
| Gunma | Maebashi | 147 | 85640 | 0.001716 | 8.6 | 2.8 | 1015.9 | 58 | 63 | 862.9 | 291.0 | 2.7 |
| Okinawa | Naha | 142 | 123860 | 0.001146 | 19.3 | 4.8 | 1018.9 | 71 | 37 | 515.1 | 304.0 | 4.3 |
| Fukui | Fukui | 122 | 71850 | 0.001698 | 8.1 | 2.8 | 1018.1 | 76 | 34 | 490.8 | 739.0 | 2.2 |
| Shiga | Otsu | 99 | 108020 | 0.000916 | 8.6 | 2.0 | — | — | — | 613.1 | 385.5 | 2.4 |
| Nara | Nara | 90 | 156510 | 0.000575 | 9.0 | 2.3 | 1018.1 | 68 | 41 | 585.2 | 383.5 | 2.5 |
| Miyagi | Sendai | 88 | 58440 | 0.001506 | 6.5 | 3.4 | 1015.8 | 66 | 48 | 664.6 | 345.0 | 1.9 |
| Niigata | Niigata | 83 | 49520 | 0.001676 | 7.1 | 3.4 | 1017.1 | 72 | 33 | 464.1 | 477.0 | 1.9 |
| Fukushima | Fukushima | 81 | 44200 | 0.001833 | 6.7 | 2.8 | 1016.2 | 66 | 45 | 623.6 | 326.5 | 2.1 |
| Nagano | Nagano | 76 | 63960 | 0.001188 | 5.0 | 2.6 | 1017.2 | 74 | 47 | 653.2 | 241.0 | 2.6 |
| Kochi | Kochi | 74 | 60710 | 0.001219 | 11.2 | 1.8 | 1017.8 | 65 | 59 | 822.8 | 601.0 | 2.8 |
| Ehime | Matsuyama | 73 | 80800 | 0.000903 | 10.5 | 2.4 | 1018.8 | 64 | 49 | 690.4 | 410.0 | 2.6 |
| Shizuoka | Shizuoka | 73 | 133080 | 0.000549 | 11.4 | 2.3 | 1015.0 | 63 | 59 | 827.7 | 719.5 | 2.6 |
| Yamagata | Yamagata | 69 | 37780 | 0.001826 | 5.0 | 1.9 | 1016.6 | 73 | 35 | 492.0 | 347.5 | 2.0 |
| Wakayama | Wakayama | 63 | 83850 | 0.000751 | 10.5 | 4.0 | 1018.1 | 63 | 48 | 682.5 | 329.0 | 2.6 |
| Oita | Oita | 60 | 63590 | 0.000944 | 10.8 | 2.8 | 1019.0 | 66 | 50 | 706.5 | 300.0 | 2.7 |
| Tochigi | Utsunomiya | 60 | 65240 | 0.000920 | 7.8 | 3.2 | 1015.7 | 65 | 59 | 806.7 | 366.5 | 2.4 |
| Yamanashi | Kofu | 57 | 85610 | 0.000666 | 8.8 | 2.4 | 1015.4 | 61 | 62 | 860.6 | 300.0 | 2.6 |
| Kumamoto | Kumamoto | 48 | 62830 | 0.000764 | 11.0 | 2.1 | 1019.3 | 70 | 47 | 665.4 | 381.5 | 2.7 |
| Saga | Saga | 47 | 61320 | 0.000766 | 10.8 | 3.2 | 1019.4 | 69 | 47 | 666.7 | 488.0 | 2.6 |
| Mie | Tsu | 45 | 86980 | 0.000517 | 9.8 | 3.5 | 1017.6 | 57 | 53 | 736.4 | 390.5 | 2.5 |
| Yamaguchi | Yamaguchi | 37 | 80260 | 0.000461 | 9.2 | 1.9 | 1019.2 | 73 | 44 | 626.4 | 552.5 | 2.4 |
| Kagawa | Takamatsu | 28 | 95660 | 0.000293 | 10.1 | 2.7 | 1018.5 | 65 | 51 | 715.5 | 309.5 | 2.6 |
| Aomori | Aomori | 27 | 39110 | 0.000690 | 3.8 | 4.0 | 1015.5 | 73 | 31 | 437.0 | 335.5 | 1.6 |
| Okayama | Okayama | 25 | 85540 | 0.000292 | 9.2 | 3.2 | 1018.6 | 67 | 50 | 707.0 | 339.0 | 2.6 |
| Shimane | Matsue | 24 | 52350 | 0.000458 | 8.8 | 3.7 | 1018.6 | 74 | 37 | 536.7 | 619.0 | 2.3 |
| Miyazaki | Miyazaki | 17 | 73410 | 0.000232 | 12.3 | 3.3 | 1018.4 | 71 | 55 | 775.6 | 407.5 | 3.1 |
| Nagasaki | Nagasaki | 17 | 80060 | 0.000212 | 11.5 | 2.4 | 1019.4 | 71 | 44 | 626.6 | 429.0 | 2.7 |
| Akita | Akita | 16 | 30610 | 0.000523 | 5.0 | 4.6 | 1016.1 | 72 | 32 | 451.5 | 506.5 | 1.7 |
| Kagoshima | Kagoshima | 10 | 48720 | 0.000205 | 13.1 | 3.3 | 1018.9 | 70 | 48 | 681.9 | 495.5 | 3.1 |
| Tokushima | Tokushima | 5 | 72850 | 0.000069 | 10.6 | 3.3 | 1018.2 | 64 | 54 | 751.4 | 309.0 | 2.8 |
| Tottori | Tottori | 3 | 62160 | 0.000048 | 8.8 | 3.4 | 1018.5 | 72 | 36 | 510.2 | 606.0 | 2.3 |
| Iwate | Morioka | 0 | 33410 | 0 | 3.3 | 3.0 | 1016.0 | 73 | 39 | 526.3 | 306.5 | 1.8 |

## Results and Conclusion

We summarized the results of the meta-regression in Table 2. A slope of the meta-regression line was significantly negative for mean air temperature (coefficient, −0.134; *P* = 0.019; Figure 1, upper panel), mean sea level air pressure (coefficient, −0.351; *P* = 0. 001; Figure 1, middle panel), and mean daily maximum UV index (coefficient, −0.908; *P* = 0.012; Figure 1, lower panel), which indicated that COVID-19 prevalence decreased significantly as air temperature, air pressure, and UV index increased. The present meta-regression suggests that temperature, pressure, and UV may be negatively associated with COVID-19 prevalence. The recent study in China has reported no association of COVID-19 epidemics with temperature and UV radiation,^2^ which, however, may be denied by the present findings of the negative association of temperature, pressure, and UV with COVID-19 prevalence in Japan.

**Table 2.** Summary of the meta-regression

| Covariate | Coefficient | Lower bound | Upper bound | *P*<br>value |
| --- | --- | --- | --- | --- |
| Mean air temperature (C) | −0.134 | −0.246 | −0.022 | 0.019 |
| Mean wind speed (m/s) | 0.042 | −0.414 | 0.498 | 0.857 |
| Mean sea level air pressure (hPa) | −0.351 | −0.562 | −0.140 | 0.001 |
| Mean relative humidity (%) | −0.042 | −0.105 | 0.021 | 0.189 |
| Mean possible sunshine (%) | −0.008 | −0.048 | 0.031 | 0.677 |
| Total sunshine duration (h) | −0.001 | −0.003 | 0.002 | 0.701 |
| Total precipitation (mm) | 0.001 | −0.002 | 0.003 | 0.679 |
| Mean daily maximum UV index | −0.908 | −1.615 | −0.201 | 0.012 |

**Figure 1.**
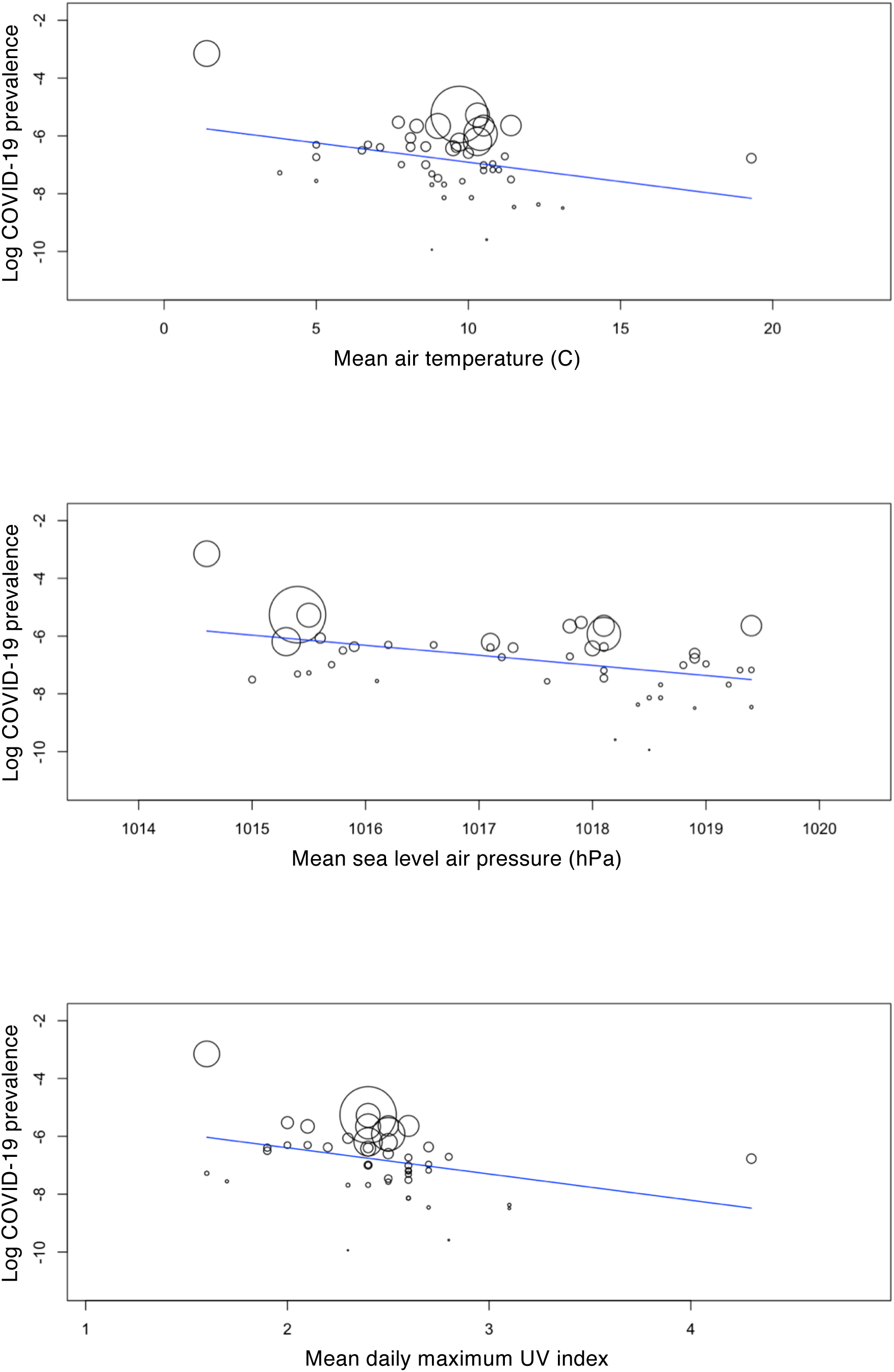
Meta-regression graph depicting the COVID-19 prevalence (plotted as the logarithm transformed prevalence on the y-axis) as a function of a given factor (plotted as a meteorological condition on the x-axis).

In conclusion, higher temperature, pressure, and UV may be associated with less COVID-19 prevalence, which should be confirmed by further epidemiological investigations taking other risk and protective factors (expect for meteorological conditions) of COVID-19 into account.

## Data Availability

The datasets generated during and/or analysed during the current study are available from the corresponding author on reasonable request.

